# Integration of molecular inflammatory interactome analyses reveals dynamics of circulating cytokines and extracellular vesicle long non-coding RNAs and mRNAs in heroin addicts during acute and protracted withdrawal

**DOI:** 10.1101/2021.06.20.21259192

**Authors:** Zunyue Zhang, Hongjin Wu, Qingyan Peng, Zhenrong Xie, Fengrong Chen, Yuru Ma, Yizhi Zhang, Yong Zhou, Jiqing Yang, Cheng Chen, Shaoyou Li, Yongjin Zhang, Yuan Wang, Yu Xu, Huayou Luo, Mei Zhu, Yiqun Kuang, Juehua Yu, Kunhua Wang

## Abstract

Heroin addiction and withdrawal influence multiple physiological functions, including immune responses, but the mechanism remains largely elusive. The objective of this study was to investigate the molecular inflammatory interactome, particularly the cytokines and transcriptome regulatory network in heroin addicts undergoing withdrawal, compared to healthy controls (HCs). Twenty-seven cytokines were simultaneously assessed in 41 heroin addicts, including 20 at the acute withdrawal (AW) stage and 21 at the protracted withdrawal (PW) stage, and 38 age- and gender-matched HCs. Disturbed T-helper(T_h_)1/T_h_2, T_h_1/T_h_17, and T_h_2/T_h_17 balances, characterized by reduced interleukin (IL)-2, elevated IL-4, IL-10, and IL-17A, but normal TNF-α, were present in the AW subjects. These imbalances were mostly restored to the baseline at the PW stage. However, the cytokines TNF-α, IL-2, IL-7, IL-10, and IL-17A remained dysregulated. This study also profiled exosomal long non-coding RNA (lncRNA) and mRNA in the plasma of heroin addicts, constructed co-expression gene regulation networks, and identified lncRNA-mRNA-pathway pairs specifically associated with alterations in cytokine profiles and T_h_1/T_h_2/T_h_17 imbalances. Altogether, a large amount of cytokine and exosomal lncRNA/mRNA expression profiling data relating to heroin withdrawal was obtained, providing a useful experimental and theoretical basis for further understanding of the pathogenic mechanisms of withdrawal symptoms in heroin addicts.

## Introduction

Globally, an estimated 40 million people with substance-use disorders (SUDs) require addiction treatment services, accounting for approximately 0.6 million drug-related deaths each year (1). To date, heroin remains one of the most abused illegal drugs in China. It is associated with a wide range of deleterious health problems and harmful economic consequences to individuals and communities (2). As an opiate drug, the administration of heroin suppresses some of the central nervous system (CNS) functioning, including heart rate, breathing, and blood pressure. The withdrawal effects are usually the opposite of the suppressive effects. In other words, instead of euphoria, reduced heart rate, and sedation, heroin addicts may experience irritable mood, anxiety, and rapid heart rate (3).

Dysregulated inflammation and increased incidence of infectious diseases like Acquired Immunodeficiency Syndrome, hepatitis, and pneumonia have been documented in heroin addicts (4). Previous studies also revealed that chronic heroin administration causes apparent suppression of some of the major components in the cellular immune system due to the presence of opioid receptors on immune cells (5, 6). Although, in the past decade, the effects of heroin and other opioids on cytokine production have been mainly investigated using cellular and animal models (7, 8), little information is available from studies on heroin addicts during withdrawal. It is noteworthy that some data from existing datasets exhibit contradictory results. For instance, a shift in T-helper(T_h_)1/T_h_2 cytokine balance toward the T_h_2 response and a suppression of T_h_1 cytokine tumor necrosis factor alpha (TNF-α) by heroin were often reported (9, 10), but some studies showed no effect or even an increase in TNF-α (11, 12). Similarly, the production of the T_h_1 signature cytokine interferon gamma (IFN-γ) could be either enhanced or suppressed by various opioids in *in vitro* studies, whereas, in animal experiments, the IFN-γ production was often diminished after exposure to opioids (10, 13). Compared with the healthy controls (HCs), the T_h_2 cytokine interleukin (IL)-4 was reported to be reduced significantly in teenage heroin addicts (14), but was also reported as unchanged in adult addicts (15). Also, the levels of the IL-10 were elevated in heroin addicts compared to controls (16). Nevertheless, it was significantly decreased in rodents within two hours after heroin administration (17). The comparison of *ex vivo* cytokine production after stimulation of whole blood between heroin users and healthy controls (HCs) revealed that heroin reduces IL-8 production, but IL-8 was reported to be significantly increased in methadone-maintained heroin addicts (12). Overall, the cytokine profiles for heroin use or abrupt cessation in individuals remain uncertain. Confounders such as age, BMI, duration of heroin exposure, or duration of withdrawal are rarely addressed.

With the advances in genome-wide sequencing and related technologies, recent studies demonstrated the influence of a systematic level regulatory network of non-coding RNAs in orchestrating a disease-specific signature (18). Long non-coding RNAs (lncRNAs) have attracted particular attention in research on the pathogenesis of various diseases, such as tumors, cardiovascular diseases, and neurodegenerative/psychiatric disorders (19). Abnormal lncRNA expression was detected not only in brain tissues but also in the peripheral blood and cerebrospinal fluid of patients with Parkinson’s disease (20). In parallel, several studies have posited that lncRNAs BACE1-AS, 17A, 51A, and NDM29 may directly or indirectly increase amyloid β protein formation and play a major role in the pathology of Alzheimer’s disease (21). Recent peripheral blood profiling studies also identified alterations in lncRNA expression patterns in patients with major depressive disorder (22, 23) and schizophrenia (24). In short, mounting evidence suggests that lncRNAs may comprise valuable diagnostic utilities and therapeutic targets in psychiatric disorders such as SUDs (25).

Exosomes are membrane-bound extracellular vesicles, usually 40–100 nm in diameter, presenting in blood, urine, saliva, cerebral spinal fluid, and breast milk (26). We and others have previously shown that the exosomal miRNAs are involved in the development of psychiatric withdrawal symptoms in patients with SUDs (27, 28), although the role of exosomal (Ex)- lncRNAs/mRNAs in SUDs remains undetermined. In addition, when evaluating the psychoneuroimmunologic factors involved in heroin addicts, there is a need to differentiate between the acute and protracted withdrawal (PW) stages. Acute withdrawal (AW) from heroin dependence induces a state of significant physiological activation and disruption of immunocompetence in heroin addicts, which seems to gradually reverse the immunological parameters to normal levels when withdrawal is sustained for an extended period (29). However, the possible mechanisms underlying molecular inflammatory interactome during heroin withdrawal are complicated and have not yet been well understood. Therefore, a careful examination of the transcriptome regulatory network, especially its impact on the immune system, is warranted.

In the present study, to investigate the molecular inflammatory interactome, particularly the cytokine associated transcriptome regulatory network in heroin addicts during withdrawal, we simultaneously profiled circulating cytokines and Ex-lncRNAs/mRNAs in plasma. We constructed gene co-expression networks based on differentially expressed Ex-lncRNAs/mRNAs in heroin addicts compared to HCs. To provide centrality between connected networks, we also identified several probable biological key drivers in the form of “hub” genes specifically associated with cytokine alterations. Consequently, our present approach uncovers the mechanism associated with the peripheral inflammatory cytokine and transcriptome regulatory network in developing withdrawal symptoms, and it pinpoints potential future therapeutic targets.

## Materials and methods

### Participants and ethics statement

A total of 79 male participants, including 41 heroin addicts currently undergoing withdrawal (n = 20 at the AW stage and n = 21 at the PW stage) and 38 healthy control subjects aged between 22 to 53, were recruited from a joint program of drug detoxification and rehabilitation in the First Affiliated Hospital of Kunming Medical University and the Kunming Drug Rehabilitation Center between January 2018 and October 2019. All protocols and recruitment procedures described in the present study were approved by the Research Ethics Committee of the First Affiliated Hospital of Kunming Medical University (2018-L-42). All participants provided written informed consent before enrollment. A clear history of heroin use and current medication was obtained via self-report and verified with caregivers and was noted in the medical record when possible. Samples were collected as previously described.(27)

### Detection of cytokine levels

In brief, fresh whole blood samples from study participants were collected into 10 mL EDTA-2Na vacuum tubes and the blood was centrifuged at 1500 g for 15 min. The plasma was transferred into a new tube and centrifuged at 20,000 g at 4°C for 15 min. The supernatant was harvested, aliquoted and stored at -80 °C until cytokine and other assays. Plasma was assessed for protein levels of 27 cytokines including bFGF (Basic Fibroblast Growth Factor), IL- 1β, IL-2, IL-4, IL-5, IL-6, IL-7, IL-8, IL-9, IL-10, IL-12p70, IL-13, IL-15, IL-17A, IP-10, IFN-γ, MCP-1 (Monocyte Chemoattractant Protein-1), MIP (Macrophage Inflammatory Protein)-1α, MIP-1β, PDGF-BB (Platelet-Derived Growth Factor-BB), RANTES (Regulated upon Activation, Normal T Cell Expressed and Presumably Secreted), TNF-α and VEGF (Vascular Endothelial Growth Factor) using a Luminex Human Magnetic Assay Kit (R&D Systems, MN, USA) according to the manufacturer’s instructions. Standard curves were generated by Bio-plex Manager software to determine unknown sample concentration.

### Exosome purification and characterization

The exosomes were isolated by size exclusion chromatography method as described previously with minor modifications (30). In brief, 2 ml of 0.8 μm-filtered plasma sample was purified using Exosupur® columns (Echobiotech, China). The exosome samples were eluted with Phosphate-buffered saline and 2 mL eluate fractions were collected. Fractions were concentrated to 200 μl by 100 kDa molecular weight cut-off Amicon® Ultra spin filters (Merck, Germany). Total RNAs containing lncRNAs from exosomes were isolated using the miRNeasy® Mini kit according to the manufacturer’s protocol.

### LncRNA and mRNA expression profiling by deep sequencing

A total amount of 1.5 μg RNA per sample was used as input material for rRNA removal using the Ribo-Zero rRNA Removal Kit (Epicentre, Madison, WI, USA). Sequencing libraries were generated using NEBNextR UltraTM Directional RNA Library Prep Kit for IlluminaR (NEB, USA) following manufacturer’s recommendations. The RNA samples were barcoded by ligation with unique adaptor sequences to allow pooling of samples. The elution containing pooled DNA library was further processed for cluster generation and sequencing using a NovaSeq 6000 platform. Library quality was analyzed using a Qubit fluorometer (Thermo Fisher Scientific, MA, USA). Raw reads were filtered using fastQC and aligned to the GRCh38 human genome assembly using HISAT2 (31). Annotations of mRNA and lncRNA in the human genome were retrieved from the GENCODE (v.25). The mRNAs and lncRNAs were quantified and analyzed using DESeq2 R package (32) and StringTie 1.3.1 (31), respectively. The number of mRNAs and lncRNAs were calculated if the fpkm >= 0.05. The mRNAs and lncRNAs with a *p*-value < 0.05 and log2 fold change > 2 were considered differentially expressed between groups.

### RT-qPCR validation

Reverse transcription and quantitative real-time PCR (RT-qPCR) was performed to validate the results of RNA-seq results. Total RNA extraction from the exosomes was performed with the miRNeasy® Mini kit (Qiagen, MD, USA) according to the manufacturer’s instructions. The concentration and quality of the RNA was determined using the RNA Nano 6000 Assay Kit of the Agilent Bioanalyzer 2100 System (Agilent Technologies, CA, USA). Single strand cDNA was synthesized with the PrimeScript™ RT reagent Kit (Takara, Dalian, China). The RT-PCRs were conducted on an ABI Prism 7500 HT using the TaqMan Fast Advanced Master Mix. Real-time PCR amplification was performed in triplicate and a negative control was included for each primer. The gene expression levels were calculated with the 2^-ΔΔCt^ method.

### Gene Ontology and KEGG enrichment analysis

Gene Ontology (GO) enrichment analysis of the differentially expressed genes (DEGs) was implemented by the clusterProfiler R packages(33). Enrichment analysis uses hypergeometric testing to find GO entries that are significantly enriched compared to the entire genome background. Meanwhile, we used clusterProfiler R packages to find KEGG pathway that are significantly enriched compared to the entire genome background.

### Statistical analysis and bioinformatics analysis

All statistical analyses were performed in R 3.6.3 Statistical Software (R Core Team, 2020). All tests were two-tailed with alpha set at p□<□0.05. The immunological data was significantly skewed and was analyzed using parametric tests. Specifically, analysis of variance (ANOVA) were implemented comparing all three groups, followed by pairwise *Student’s tests*. Pearson’s correlation coefficients and Spearman’s rank correlation were implemented to analyze the correlations between heroin addicts’ clinical characteristics and cytokines. Statistical analysis and significance were calculated using various tests and adjusted for multiple testing. Sequencing data have been deposited in the Gene Expression Omnibus database under GSE172306.

## Results

### Clinical characteristics

This study recruited 41 male heroin addicts, including 20 AW subjects at 7∼14 days after the initiation of abstinence and 21 protracted PW subjects who had abstained from heron for approximately one year, and 38 age-matched healthy male volunteers. The main clinical characteristics of the study participants are summarized in **Table 1**. Although heroin was reported as the primary abused substance and all individuals in the two addicted groups had a diagnosis of Opioid Use Disorder, 25% of AW subjects (n = 5) and 28.6% of PW subjects (n = 6) were also methamphetamine users. Also, 45% of AW subjects (n = 9) and 61.9% of PW subjects (n = 13) used drugs via injection, whereas 50% of AW subjects (n = 10) and 33.3% of PW subjects (n = 7) used drugs via snorting. The others (n = 1 per group) used both routes. There were no significant differences for any variables, including age, BMI, substance-use history, and education level, between the two groups of heroin addicts and HCs.

**Table 1.** Demographic and Clinical Characteristics of all participants. Age-matched healthy male volunteers (HC, n=38) and heroin addicts including acute withdrawal (AW, n=20) subjects with 7∼14 days after initiation of abstinence and protracted withdrawal (PW, n=21) subjects who had abstained from heron for approximately 1 year were recruited in this study. Note: Values are showed as mean ± SD, medians (interquartile ranges) or numbers of participants (%). BMI, Body Mass Index; NA, Not Applicable.

|  | Healthy<br>Control<br>(n=38) | Acute<br>Withdrawal<br>(n=20) | Prostrated<br>Withdrawal<br>(n=21) | *p* |
| --- | --- | --- | --- | --- |
| Age (year) | 37.32±8.29 | 37.7±7.30 | 37.62±8.7 | 0.7502 |
| Marriage |  |  |  |  |
| Married | 15(39.5%) | 8(40.0%) | 9(42.9%) | 1 |
| Unmarried | 11(28.9%) | 6(30.0%) | 6(28.6%) |  |
| Divorced | 12(31.6%) | 6(30.0%) | 6(28.6%) |  |
| Education |  |  |  |  |
| Illiteracy | 6(15.8%) | 4(20.0%) | 3(14.3%) | 0.9597 |
| Primary school | 14(36.8%) | 7(35.0%) | 7(33.3%) |  |
| Junior high school | 10(26.3) | 6(30.0%) | 9(42.9%) |  |
| High school | 6(15.8) | 2(10/0%) | 2(9.5%) |  |
| College | 2(5.3%) | 1(5.0%) | 0 |  |
| Height (cm) | 165.84±7.38 | 166.65±6.95 | 166.7±6.76 | 0.8702 |
| Weight (kg) | 60.76±9.23 | 60.34±8.97 | 60.24±7.77 | 0.9634 |
| BMI | 22.03 | 21.64 | 21.63 | 0.7513 |
| Drug type |  |  |  |  |
| Heroin | NA | 15(75.0%) | 15(71.4%) | 1 |
| Heroin & Meth | NA | 5(25.0%) | 6(28.6%) |  |
| Route |  |  |  |  |
| injection | NA | 9(45.0%) | 13(61.9%) | 0.6623 |
| snorting | NA | 10(50.0%) | 7(33.3%) |  |
| both |  | 1(5.0%) | 1(4.8%) |  |
| Drug history (year) | NA | 7.25±3.8 | 7.2±3.39 | 0.8859 |

### Alterations in cytokine profiles in heroin addicts during withdrawal

In the present study, to investigate the alterations of circulating cytokine associated with heroin addiction and withdrawal, we analyzed the concentrations of traditional cytokines in the peripheral blood from heroin addicts and age-matched HCs. Blood samples were collected, processed, and investigated using Luminex Human Cytokine 27-plex assay (**Materials and methods**). In the cytokines measured, the INF-γ and GM-CSF fell below the lower level of quantitation in > 20% of samples and were excluded from analysis. The analysis revealed that the levels of IL-1ra, IL-1β, IL-5, IL-12p70, IL-13, IL-15, MIP-1β, bFGF, and RANTES in the plasma of heroin addicts in the AW and PW stages were not significantly different from those in the plasma of the control group subjects (data not shown). Significantly lower levels of IL-2 (*adj*.*p* = 2.78E-03) and PDGF-BB (*adj*.*p* = 2.01E-05) were observed in AW subjects compared with those in HCs (**Table 2**). Simultaneously, significantly higher levels of IL-4 (*adj*.*p* = 1.59E- 04), IL-10 (*adj*.*p* = 8.74 E-06), G-CSF (*adj*.*p* = 8.75E-09), Eotaxin (*adj*.*p* = 4.31E-09), MCP-1 (*adj*.*p* = 1.61E-05), IL-17A (*adj*.*p* = 3.85E-03), and VEGF (*adj*.*p* = 7.64E-07) were found in AW subjects than in the HCs (**Table 2**). A similar analysis was conducted for cytokine profiles in PW heroin addicts. Compared to HCs, there were significant fluctuations in the levels of IL-2 (*adj*.*p* = 0.047), IL-7 (*adj*.*p* = 1.58E-04), IL-10 (*adj*.*p* = 0.047), IL-17A (*adj*.*p* = 5.41E-03), MIP-1α (*adj*.*p* = 8.93E-03), and TNF-α (*adj*.*p* = 3.45E-03) in PW subjects (**Table 2**). We next compared the cytokine profiles of PW subjects with those of AW subjects. Significant differences in IL-2 (*adj*.*p* = 4.48E-03), IL-4 (*adj*.*p* = 1.59E-09), IL-10 (*adj*.*p* = 8.74E-06), IL-17A (*adj*.*p* = 3.85E-03), G-CSF (*adj*.*p* = 8.75E-09), Eotaxin (*adj*.*p* = 4.31E-09), MCP-1 (*adj*.*p* = 1.61E-05), MIP-1α (*adj*.*p* = 8.93E-03), PDGF-BB (*adj*.*p* = 7.25E-04), TNF-α (*adj*.*p* = 6.62E-4), and VEGF (*adj*.*p* = 7.64E-07) were detected (**Table 2**), suggesting that the majority of heroin acute discontinuation induced cytokine abnormalities (e.g., IL-4, Eotaxin, G-CSF, MCP-1, PDGF-BB, and VEGF) could be restored to normal range after a certain period of withdrawal (**Fig. 1A-E**). However, several cytokines, IL-2, IL-7, IL-10, and IL-17A were dramatically altered at the AW stage, and could be only partially restored even a year after the initiation of abstinence (**Fig. 1E-I**). In addition, it was noteworthy that the TNF-α and MIP-1α were only dysregulated in PW heroin addicts, not at the AW stage, but within 7∼14 days after initiating abstinence (**Fig. 1J**).

**Table 2.** Alterations of cytokine levels in heroin addicts (AW and PW) versus healthy controls (HC). Note: Values are showed as mean ± SD.

|  | mean | mean | mean | Healthy control vs Heroin AW |  |  | Healthy control vs Heroin PW |  |  | Heroin PW vs AW |  |  |
| --- | --- | --- | --- | --- | --- | --- | --- | --- | --- | --- | --- | --- |
|  | Healthy Control | Heroin AW | Heroin PW | Fold change | *p.value* | *adj.p* | Fold change | *p.value* | *adj.p* | Fold change | *p.value* | *adj.p* |
| IL-2 | 7.02±2.86 | 2.57±4.93 | 14.17±15.30 | 2.74 | **9.26E-04** | **2.78E-03** | 0.50 | **0.0467** | **0.0467** | 0.18 | **2.98E-03** | **4.48E-03** |
| IL-4 | 3.58±1.43 | 15.56±4.80 | 4.86±2.82 | 0.23 | **4.58E-10** | **1.38E-09** | 0.74 | 0.06 | 0.06 | 3.20 | **1.05E-09** | **1.59E-09** |
| IL-5 | 13.85±11.10 | 17.90±28.34 | 21.71±25.27 | 0.77 | 0.55 | 0.65 | 0.64 | 0.19 | 0.56 | 0.82 | 0.65 | 0.65 |
| IL-6 | 2.16±1.66 | 6.71±6.12 | 6.85±16.04 | 0.32 | **3.77E-03** | **0.01** | 0.31 | 0.20 | 0.29 | 0.98 | 0.97 | 0.97 |
| IL-7 | 82.10±27.61 | 236.96±103.74 | 188.34±100.88 | 0.35 | **1.96E-06** | **5.89E-06** | 0.44 | **1.05E-04** | **1.58E-04** | 1.26 | 0.14 | 0.14 |
| IL-8 | 21.67±12.07 | 21.63±11.37 | 29.89±13.75 | 1.00 | 0.99 | 0.99 | 0.73 | **0.03** | 0.06 | 0.72 | **0.04** | 0.06 |
| IL-10 | 5.15±4.11 | 39.11±15.41 | 12.56±15.31 | 0.13 | **2.97E-08** | **8.91E-08** | 0.41 | **0.0461** | **0.0461** | 3.12 | **5.83E-06** | **8.74E-06** |
| IL-13 | 7.26±8.53 | 5.81±5.41 | 6.83±6.04 | 1.25 | 0.43 | 0.82 | 1.06 | 0.82 | 0.82 | 0.85 | 0.57 | 0.82 |
| IL-17A | 14.03±6.97 | 37.17±17.67 | 22.02±111.03 | 0.38 | **1.12E-05** | **3.37E-05** | 0.64 | **5.41E-03** | **5.41E-03** | 1.69 | **2.56E-03** | **3.85E-03** |
| Basic FGF | 261.77±140.75 | 257.16±75.53 | 302.48±144.97 | 1.02 | 0.87 | 0.87 | 0.87 | 0.30 | 0.45 | 0.85 | 0.22 | 0.45 |
| G-CSF | 254.41±92.42 | 839.87±264.43 | 290.23±107.94 | 0.30 | **3.19E-09** | **8.75E-09** | 0.88 | 0.21 | 0.21 | 2.89 | **5.83E-09** | **8.75E-09** |
| Eotaxin | 169.89±85.67 | 914.96±329.08 | 149.04±57.95 | 0.19 | **2.87E-09** | **4.31E-09** | 1.14 | 0.27 | 0.27 | 6.14 | **1.93E-09** | **4.31E-09** |
| MCP-1 | 63.13±30.94 | 94.41±30.14 | 49.02±23.99 | 0.66 | **4.30E-04** | **6.45E-04** | 1.27 | 0.08 | 0.08 | 1.93 | **5.52E-06** | **1.66E-05** |
| MIP-1α | 4.02±3.05 | 4.37±1.21 | 6.39±2.84 | 0.92 | 0.54 | 0.54 | 0.63 | **4.59E-03** | **8.93E-03** | 0.68 | **5.95E-03** | **8.93E-03** |
| PDGF-BB | 10095.47±8850.47 | 2440.12±1814.73 | 10243.75±8533.97 | 4.14 | **6.71E-06** | **2.01E-05** | 0.99 | 0.95 | 0.95 | 0.24 | **4.83E-04** | **7.25E-04** |
| TNFα | 77.91±18.43 | 69.50±18.56 | 104.23±33.11 | 1.12 | 0.11 | 0.11 | 0.75 | **2.30E-03** | **3.45E-03** | 0.67 | **2.21E-04** | **6.62E-04** |
| VEGF | 578.83±547.43 | 1244.71±498.93 | 405.09±241.34 | 0.47 | **3.81E-05** | **5.71E-05** | 1.43 | 0.11 | 0.11 | 3.07 | **2.55E-07** | **7.64E-07** |

**Figure 1.**
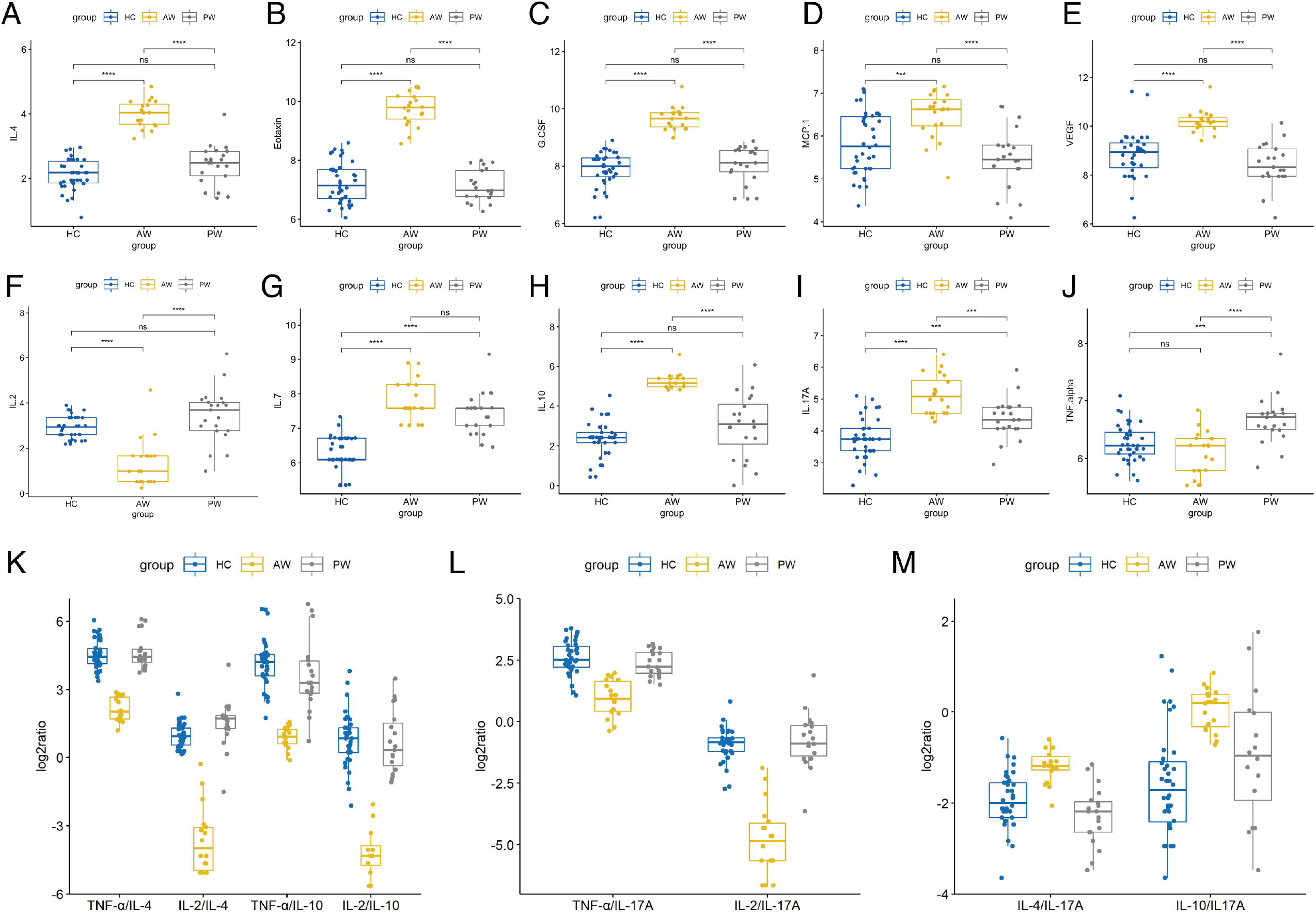
Alterations in cytokine profiles in heroin addicts during withdrawal. A-J. The levels of various cytokines in the plasma samples of healthy controls, heroin addicts from AW and PW groups was investigated, including IL-4 (A), Eotaxin (B), G-CSF (C), MCP-1 (D), VEGF (E), IL-2 (F), IL-7 (G), IL-10 (H), IL-17A (I), TNF-alpha (J). K-M. Th1/Th2/Th17/Treg balance in heroin addicts across two withdrawal stages (AW and PW). The ratio of T_h_1/T_h_2, T_h_1/T_reg_ (K) and T_h_1/T_h_17 (L) was significantly decreased in AW stage, and recovered in PW stage. The ratio of T_h_2/T_h_17 and T_reg_/T_h_17 (M) was significantly increased in AW stage, and recovered in PW stage. Each dot represents one patient in each group. All intergroup comparisons by Mann-Whitney U-test.

We then sought to examine the T_h_1/T_h_2/T_h_17 balance in heroin addicts across two withdrawal stages. We computed the T_h_1/T_h_2, T_h_1/T_h_17, and T_h_2/T_h_17 using the ratios among the following three categories of cytokines, T_h_1 (TNF-α or IL-2), T_h_2 (IL-4 or IL-10), and T_h_17 (IL- 17A). Notably, the ratio of T_h_1/T_h_2 and T_h_1/T_h_17 was significantly decreased in AW subjects, relative to PW heroin addicts and HCs (**Fig. 1K-L**). Conversely, the ratio of T_h_2/T_h_17 was significantly increased in heroin addicts at the AW stage, compared to the subjects in the other two groups (**Fig. 1M**). Taken together, the analyses of cytokine profiles indicate a dramatic disturbance in the immune system function and a shift in T_h_1/T_h_2/T_h_17 balance at the stage of acute discontinuation and a partial restoration after long-term withdrawal (**Table 3**).

**Table 3.** T_h_1/T_h_2/T_h_17 balance in heroin addicts across two withdrawal stages (AW and PW).

|  |  | mean | mean | mean | Healthy control vs Heroin AW |  | Healthy control vs Heroin PW |  | Heroin PW vs AW |  |
| --- | --- | --- | --- | --- | --- | --- | --- | --- | --- | --- |
|  |  | Healthy Control | Heroin AW | Heroin PW | *p.value* | *adj.p* | *p.value* | *adj.p* | *p.value* | *adj.p* |
| Th1/Th2 | TNF-α/IL-4 | 25.52±12.28 | 4.63±1.68 | 28.93±17.83 | **8.92E-13** | **2.68E-12** | 0.4599 | 0.6899 | **1.25E-05** | **3.78E-05** |
| | IL-2/IL-4 | 2.16±1.04 | 0.14±0.2 | 3.67±3.44 | **1.24E-14** | **1.18E-13** | 0.0753 | 0.2260 | **2.91E-04** | **6.56E-04** |
| Th1/Th17 | TNF-α/IL-17A | 6.7±3.05 | 2.15±1.04 | 5.48±1.96 | **6.01E-11** | **1.35E-10** | 0.0718 | 0.2260 | **5.29E-07** | **2.38E-06** |
| | IL-2/IL-17A | 0.57±0.28 | 0.06±0.07 | 0.78±0.78 | **3.45E-14** | **1.55E-13** | 0.2706 | 0.4871 | **7.43E-04** | **1.34E-03** |
| Th1/Treg | TNF-α/IL-10 | 34.69±56.64 | 1.92±0.57 | 31.89±48.08 | **1.02E-03** | **1.02E-03** | 0.8460 | 0.8763 | **1.42E-02** | **1.58E-02** |
| | IL-2/IL-10 | 2.43±2.64 | 0.05±0.06 | 2.55±2.84 | **2.54E-06** | **4.57E-06** | 0.8763 | 0.8763 | **1.22E-03** | **1.84E-03** |
| Th2/Th17 | IL-4/IL-17A | 0.28±0.12 | 0.45±0.11 | 0.22±0.10 | **5.58E-06** | **8.37E-06** | 0.0524 | 0.2260 | **9.65E-08** | **8.68E-07** |
| Th2/Treg | IL-4/IL-10 | 1.19±1.20 | 0.43±0.09 | 0.82±0.76 | **4.31E-04** | **4.85E-04** | 0.1607 | 0.3615 | **4.16E-02** | **4.16E-02** |
| Th17/Treg | IL-17A/IL-10 | 4.92±5.65 | 1.01±0.33 | 5.42±6.44 | **1.36E-04** | **1.75E-04** | 0.7742 | 0.8763 | **7.91E-03** | **1.02E-02** |

We further analyzed the correlation between clinical characteristics (age, BMI, etc.) and the above dysregulated cytokines identified in heroin addicts. As shown in **Supplementary Table. 1**, bFGF (*ρ* = -0.34, *p* = 0.0235) was significantly negatively correlated with the type of drug. IL-8 (*ρ* = 0.34, *p* = 0.0387), bFGF (*ρ* = 0.42, *p* = 0.0085), MIP-1α (*ρ* = 0.34, *p* = 0.0001), and PDGF-BB (*ρ* = 0.36, *p* = 0.026) were significantly positively correlated with the routes of heroin administration. IL-7 (*ρ* = 0.36, *p* = 0.028), IL-13 (*ρ* = 0.32, *p* = 0.048), bFGF (*ρ* = 0.39, *p* = 0.0165), and PDGF-BB (*ρ* = 0.40, *p* = 0.0147) were significantly positively correlated with education. IL-7 is important in the homeostatic maintenance and peripheral expansion of naïve T cells. It was significantly linearly negatively correlated with age (*γ* = -0.34, *p* = 0.0407), in line with the previous report showing that IL-7 might play a role in age-related changes. (34) Furthermore, the ratios of T_h_1/T_h_17 and T_h_2/T_h_17 in the heroin addicts did not correlate with any variable, while the ratios of T_h_1/T_h_2 (TNF-α/IL-10, *γ* = -0.34, *p* = 0.0401, and IL-2/IL-10, *γ* = -0.33, *p* = 0.0434) were linearly correlated with the years of drug-intake (**Supplementary Table 1**).

### Differential expression analysis of Ex-mRNAs and Ex-lncRNAs between heroin addicts and healthy controls

To gain insight into the transcriptional dynamics of extracellular vesicles, we utilized the size exclusion chromatography approach to isolate exosomes derived from the peripheral blood of selected heroin addicts (n = 20) and HCs (n = 10) for RNA sequencing.(27) Exosomes were characterized and verified using transmission electron microscopy, nanoparticle tracking analysis, and western blotting assays. CD63 and CD81 served as exosome surface markers, while Calnexin was used as a negative marker, and their expression patterns were verified by western blot results (**Supplementary Fig. 1**).

Next, the total RNA of plasma exosomes extracted from heroin addicts and HCs were subjected to RNA sequencing using the Illumina NovaSeq platform. A total of 94, 361 transcripts were assembled, including 25,932 mRNA (19,986 known and 5,946 new genes) and 6,8429 lncRNA transcripts (32,485 known and 35,944 putative new lncRNAs), which were then eventually filtered down to 12,158 mRNA and 10,297 lncRNA transcripts for final analysis. A total of 185 differentially expressed Ex-mRNAs exhibited a more than two-fold change, *p* < 0.05, of which 108 were upregulated, and 77 were downregulated comparing the AW and HC groups; they are indicated in an MA plot and a volcano plot (**Fig. 2A**,**D**). In the comparison of the PW group and the HCs, a total of 186 differentially expressed Ex-mRNAs were identified, of which 57 were upregulated, and 129 were downregulated (**Fig. 2B**,**E**). When comparing PW and AW, we identified a total of 181 differentially expressed Ex-mRNAs, of which 75 were upregulated, and 106 were downregulated (**Fig. 2C**,**F**). The differentially expressed Ex-mRNAs are shown in heatmaps (**Fig. 2G-I**).

**Figure 2.**
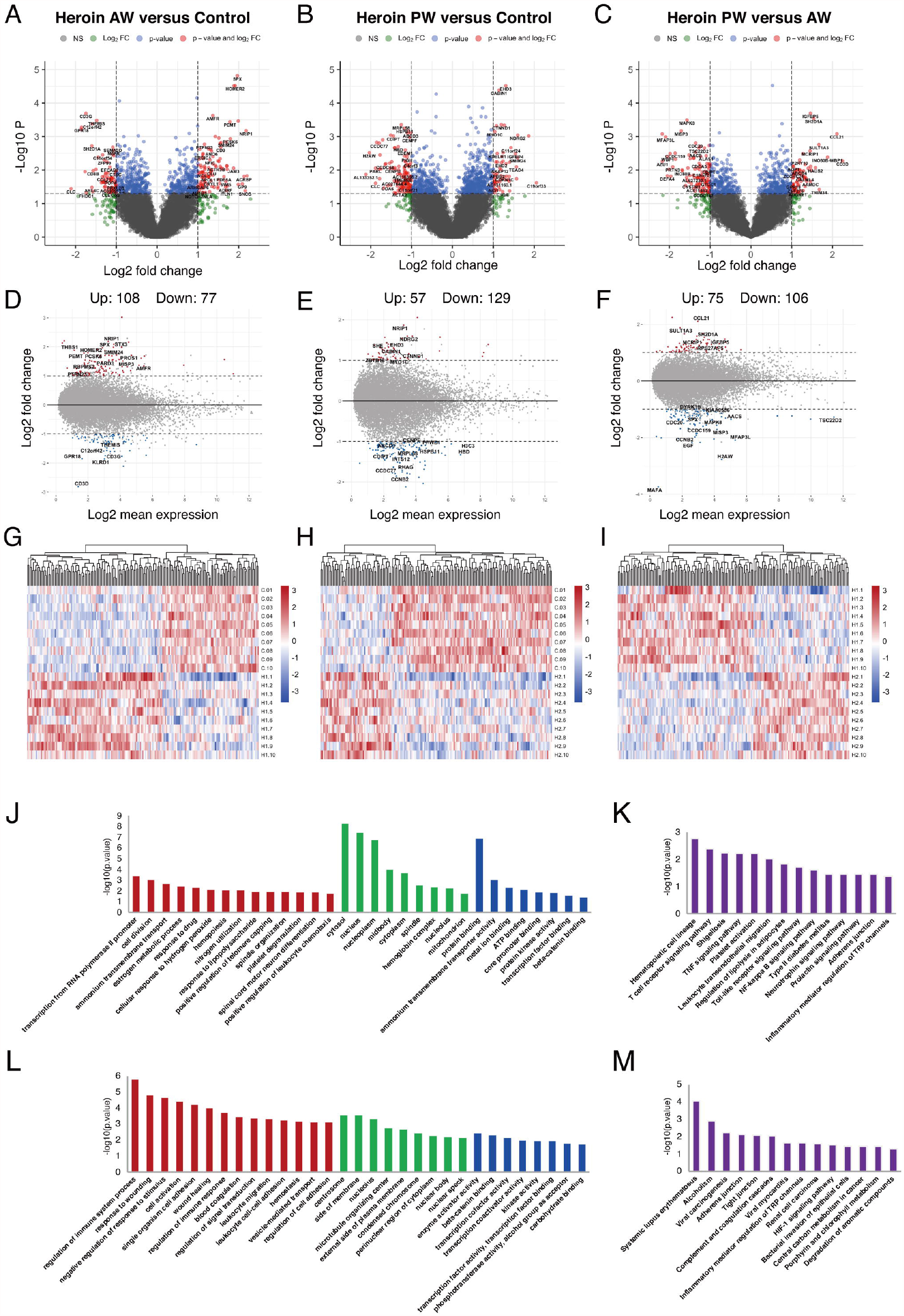
Identification and functional enrichment analysis of differentially expressed Ex-mRNA. A-B. The volcano plot (A) and hierarchical clustering heatmap (B) of differentially expressed exosome mRNAs in Heroin AW versus HC (P < 0.05, fold change > 1.5). C-D. The volcano plot (C) and hierarchical clustering heatmap (D) of differentially expressed exosome mRNAs in Heroin PW versus HC (P < 0.05, fold change > 1.5). E-F. The volcano plot (E) and hierarchical clustering heatmap (F) of differentially expressed exosome mRNAs in Heroin AW versus Heroin PW (P < 0.05, fold change > 1.5). G-H. Top enrichment of KEGG pathways related to differentially expressed exosome mRNAs.

Similar analyses were performed for the Ex-lncRNAs, and we identified 231 lncRNA transcripts exhibiting a more than two-fold change, *p* < 0.05, 123 of which were upregulated, and 108 were downregulated when comparing the AW group and the HCs (**Fig. 3A**,**D**). A comparison of PW group and the HCs shows that we identified 375 Ex-lncRNA differentially regulated transcripts, of which 143 were upregulated, and 232 were downregulated (**Fig. 3B**,**E**). Moreover, when comparing PW and AW, we identified 365 differentially expressed Ex-lncRNA transcripts, of which 143 were upregulated, and 222 were downregulated (**Fig. 3C**,**F**). These differentially expressed Ex-lncRNAs are shown in heatmaps (**Fig. 3G-I**). Correspondingly, the fold changes of the 20 most significant differentially expressed EV-mRNAs and Ex-lncRNA are listed in **Supplementary Tables 2 & 3**, respectively.

**Figure 3.**
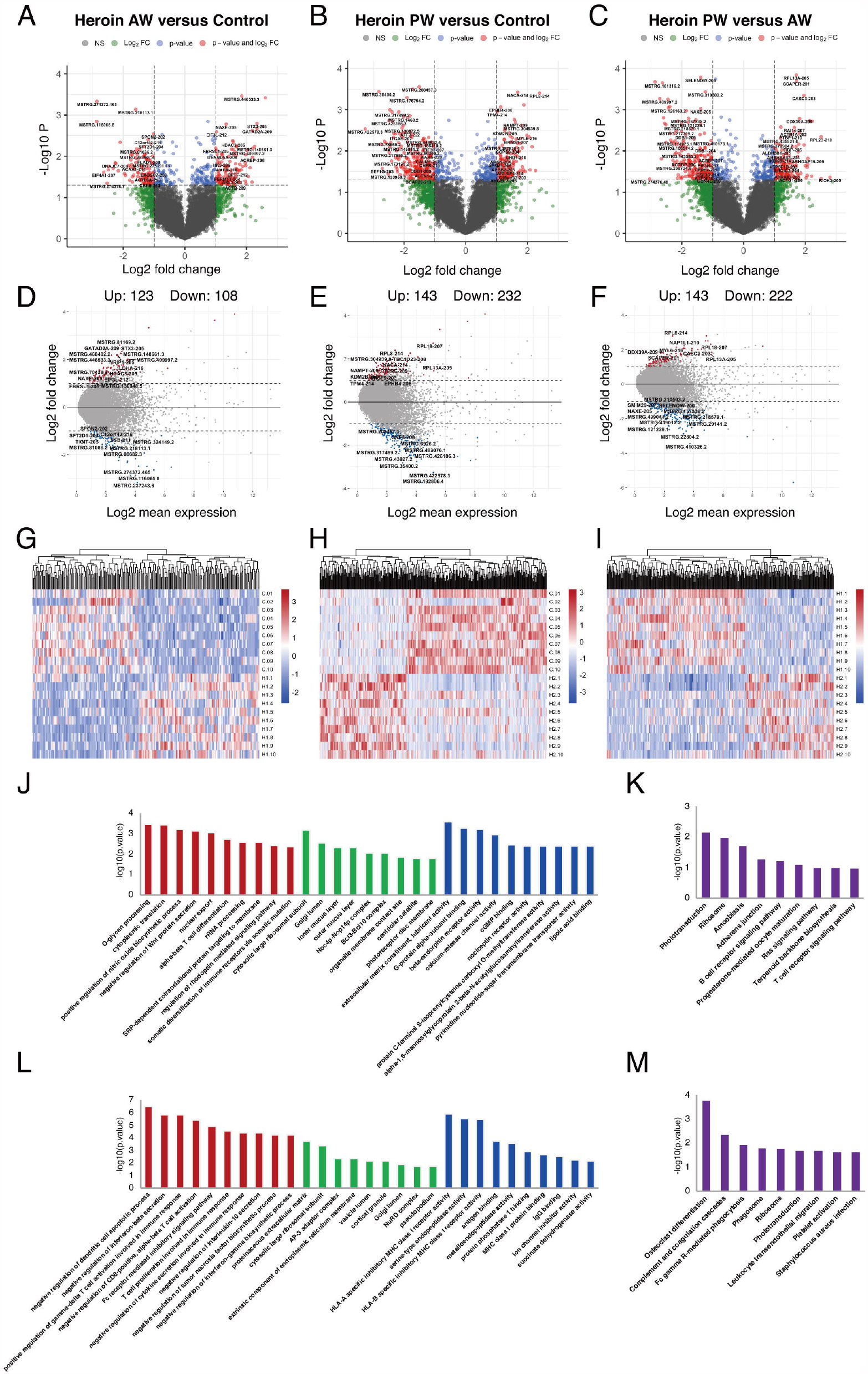
Identification and functional enrichment analysis of differentially expressed exosome lncRNAs. A-B. The volcano plot (A) and hierarchical clustering heatmap (B) of differentially expressed exosome lncRNAs in Heroin AW versus HC (P < 0.05, fold change > 1.5). C-D. The volcano plot (C) and hierarchical clustering heatmap (D) of differentially expressed exosome lncRNAs in Heroin PW versus HC (P < 0.05, fold change > 1.5). E-F. The volcano plot (E) and hierarchical clustering heatmap (F) of differentially expressed exosome lncRNAs in Heroin AW versus Heroin PW (P < 0.05, fold change > 1.5). G-H. Top enrichment of KEGG pathways related to differentially expressed exosome lncRNAs.

### Functional annotation and identification of differentially expressed Ex-mRNAs and Ex-lncRNAs

GO and KEGG pathway enrichment analyses were conducted to gain insight into the biological characteristics of the differentially expressed Ex-mRNAs. When comparing AW subjects to HCs, the GO terms associated with differentially expressed Ex-mRNAs are shown in **Fig. 3 J–K**: For the biological process, the differentially expressed Ex-mRNAs appeared to be most significantly enriched in *regulation of immune system process* (*p* = 1.64E-06), and *response to wounding* (*p* = 1.56E-05); for the cellular component, the most significantly enriched term was *centrosome* (*p* = 2.72E-04); and for the molecular function, the most significantly enriched term was *enzyme activator activity* (*p* = 0.0038) and *β-catenin binding* (*p* = 0.0047). The KEGG results indicated that differentially expressed Ex-mRNAs between the AW group and HCs were involved in *Hematopoietic cell lineage* (*p* = 0.0017), *T cell receptor signaling pathway* (*p* = 0.004), *Shigellosis* (*p* = 0.0058), and *TNF signaling pathway* (*p* = 0.0061).

As is shown in **Fig. 2 L-M**, the same analyses were applied to the differentially expressed Ex-mRNAs for the PW group and the HCs: For the biological process, the most significantly enriched term was transcription from RNA polymerase II promoter (*p* = 4.06E-04) and cell division (*p* = 9.14E-04); for the cellular component, the most significantly enriched term was cytosol (*p* = 4.97E-09); and for the molecular function, the most significantly enriched term was protein binding (*p* = 1.23E-07). Moreover, the KEGG analysis indicated that differentially expressed Ex-mRNAs in the PW group compared to the HCs were involved in *Systemic lupus erythematosus* (*p* = 8.60E-05) and *Alcoholism* (*p* = 0.0012).

GO and KEGG pathway enrichment analyses were conducted to investigate the potential function of genes related to differentially expressed Ex-lncRNAs. When comparing AW to HCs, the enriched GO terms associated with genes related to differentially expressed Ex-lncRNAs are shown in **Fig. 3 J-M**: For the biological process, the most significantly enriched term was *O-glycan processing* (*p* = 3.40E-04) and *cytoplasmic translation* (*p* = 3.71E-04); for the cellular component, the most significantly enriched term was *cytosolic large ribosomal subunit* (*p* = 6.74E-04); and for the molecular function, the most significantly enriched term was *extracellular matrix constituent, lubricant activity* (*p* = 2.63E-04, **Fig. 3J**). The KEGG results indicated that genes related to differentially expressed Ex-lncRNAs between AW and HCs were mainly involved in *Phototransduction* (*p* = 0.0067) and *Ribosome* (*p* = 0.0099, **Fig. 3K**).

The same analyses were applied to the differentially expressed Ex-lncRNAs between the PW group and the HCs (**Fig. 3 L-M**): For the biological process, the majority of differentially expressed Ex-lncRNAs appeared to be associated with *negative regulation of dendritic cell apoptotic process* (*p* = 3.15E-07), *negative regulation of interferon-β secretion* (*p* = 1.45E-06), and *positive regulation of γ-δ T cell activation involved in immune response* (*p* = 1.45E-06); for the cellular component, the most significantly enriched term was *proteinaceous extracellular matrix* (*p* = 1.76E-04); and for the molecular function, the most significantly enriched term was *HLA-A specific inhibitory MHC class I receptor activity* (*p* = 1.39E-06) and *serine-type endopeptidase activity* (*p* = 3.13E-06). Furthermore, the KEGG results indicated that differentially expressed Ex-lncRNAs in PW and HCs were significantly involved in *Osteoclast differentiation* (*p* = 1.65E-04), *Complement and coagulation cascades* (*p* = 0.004), and *Fc γ R-mediated phagocytosis* (*p* = 0.011). Altogether, these analyses indicate that the unique pattern of altered immune-related signaling pathways in AW and PW represents the pathogenesis of disrupted immunocompetence associated with heroin withdrawal stages.

### Correlation analysis of differential expressed Ex-lncRNAs/mRNAs and cytokines

A transcriptome regulatory network was constructed using a combination of Ex-lncRNAs and Ex-mRNAs differentially expressed across various comparisons to explore the association and possible mechanism of inflammatory interactome between exosomal transcriptome and cytokine profiles during heroin withdrawal. We implemented a dynamic tree-cutting algorithm with a robust measure of pairwise interconnectedness in the WGCNA R package to construct a correlation network. We then created a cluster tree and defined modules as branches.(35) WGCNA analysis with a soft-thresholding power value equal to 7 revealed the co-expression of genes/transcripts in a total of five modules. Each module was assigned a unique color label: blue, turquoise, green, red, and brown, as shown under the cluster dendrogram (**Fig. 4A**). The matrix with the module-trait relationships and corresponding *p* values between these five identified modules on group traits are shown in a heatmap (**Fig. 4B**). The Module eigengenes (ME) were calculated and could represent each module. Among the five modules, the MEs of two modules MEblue (*r* = 0.79, *p*□=□2.0E-11) and MEred (*r* = 0.87, *p*□=□9.0E-10) were positively correlated with the heroin addicts in the AW group, comprising of Ex-lncRNAs/Ex-mRNAs with relatively higher expression levels in the AW group compared to the control and PW groups. In another module, turquoise, the MEturquoise was positively correlated with the heroin addicts in the PW group (*r* = 0.9, *p*□=□2.0E-11) and comprised transcripts with higher expression levels in the heroin addicts in the PW group relative to the other two groups. In contrast, in the other two modules, the MEgreen (*r* = -0.81, *p*□=□1.0E-7) and MEbrown (*r* = -0.8, *p*□=□2.0E-7) were correlated inversely with the AW and PW groups, respectively.

**Fig. 4.**
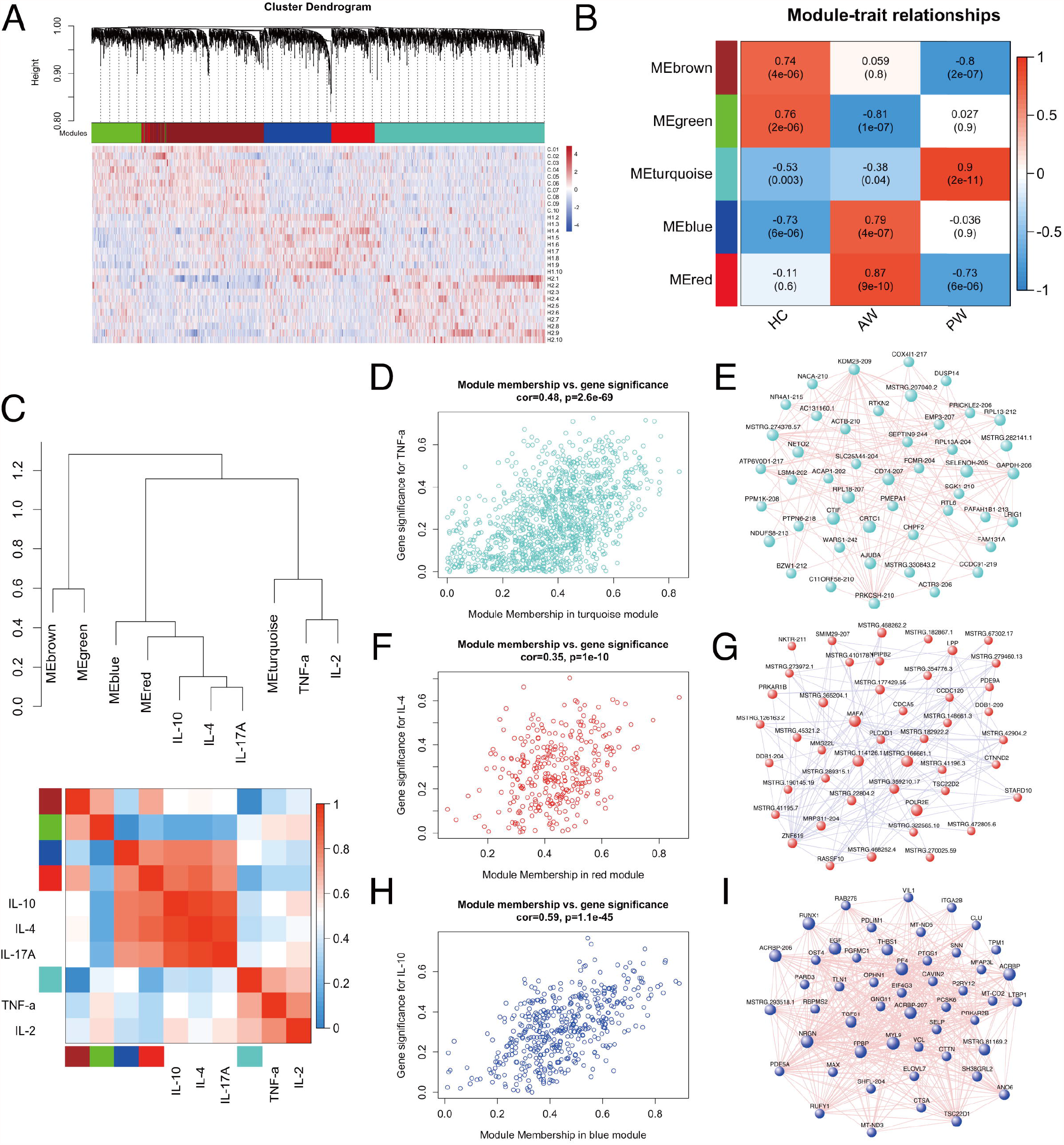
WGCNA of plasma exosome samples uncovered dynamic changes of exosome mRNAs and lncRNAs signatures during substance withdrawal. (A) Hierarchical cluster dendrogram using WGCNA analyzing RNA sequencing data. A total of 5 modules were identified after 0.25 threshold merging. (B) Module trait correlation analysis revealed that 5 key modules were significantly correlated with Heroin AW and Heroin PW. (C) The eigengene dendrogram and heatmap showed the relevancies of cytokines and identified RNA-seq modules. TNF-*α*, IL-2, IL-4, IL-10 and IL-17A were selected. (D-F) The correlation and connectivity of the turquoise module and TNF-*α* (*r* = 0.48, *p* = 2.6E-69) (D), the red module and IL-4 (*r* = 0.35, *p* = 1.1E-10) (E), the blue module and IL-10 (*r* = 0.59, *p* = 1.1E-45) (F). G-I. The hub lncRNAs/mRNAs of the turquoise (G), red (H) and blue (I) modules.

Next, we selected the TNF-*α*, IL-2, IL-4, IL-10, and IL-17A to determine their relevancies to those identified modules. As shown in **Fig. 4C**, the eigengene dendrogram and heatmap indicated that the turquoise module was clustered with TNF-*α* and IL-2, whereas the red and blue modules were clustered with IL4, IL-10, and IL-17A, which were also supported by gene significance and module membership assays. The turquoise, red, and blue modules were selected for subsequent analyses. The turquoise module, which was comprised of 1,185 differentially expressed genes/transcripts including 497 Ex-lncRNAs and 688 Ex-mRNAs, displayed a strong correlation between gene significance for TNF-*α* and connectivity (*r* = 0.48, *p* = 2.6E-69, **Fig. 4D**). The genes/transcripts in the turquoise module were ranked by their degree of gene co-expression connectivity, and the 50 genes with the highest connectivity were classified as candidate hub genes for further analysis, including CD74, major histocompatibility complex, class I, E (HLA-E), SELENOH-205, and RPL18-207 (**Fig. 4E**). These genes were highly expressed in the heroin addicts of the PW group relative to the other two groups. Similar analyses were conducted on the red and blue modules. The blue (473 genes) and the red (322 genes) modules showed strong correlations between gene significance for IL-4 (*r* = 0.35, *p* = 1.1E-10, **Fig. 4F**) and IL-10 (*r* = 0.59, *p* = 1.1E-45, **Fig. 4H**) and their connectivity, respectively. Hub lncRNAs/mRNAs were then identified in the red module, including RNA polymerase II, I and III subunit E (POLR2E), MSTRG.462778.1, MSTRG.114126.1, and MSTRG.166661.1 (**Fig. 4G**), and in the blue module, including pro-platelet basic protein (PPBP), platelet factor 4 (PF4), myosin light chain 9 (MYL9), prostaglandin-endoperoxide synthase 1 (PTGS1) and MSTRG.293518.1 (**Fig. 4I**). These hub lncRNAs/mRNAs of the red and blue modules were significantly upregulated in the AW group of heroin addicts compared to the control and PW groups.

These hub lncRNAs/mRNAs were analyzed for their expression by qPCR. It was found that the seven lncRNAs/mRNAs from modules blue and red were significantly upregulated in the AW group of heroin addicts compared to the two other groups. The three lncRNAs/mRNAs from the turquoise module were significantly upregulated in the PW subjects compared to those from the other two groups (**Supplementary Table 2**), consistent with the observations from RNA-seq analysis. However, one lncRNA in the red module (MSTRG.114126.1) was upregulated in AW heroin addicts in RNA-seq analysis, but it was not statistically significant (**Supplementary Table 3**). To gain an insight into the function of immune-related lncRNAs and mRNA, we further examined the lncRNA/mRNA-pathway pairs identified in each module. We identified a lncRNA/mRNA-pathway regulatory network. This regulatory network involves 17 lncRNA, 43 mRNA, and 11 immune-related pathways. The majority of Ex-lncRNA/mRNA from red and blue modules and their targeted mRNAs were correlated with *Platelet activation, Chemokine signaling pathway*, and *Leukocyte trans-endothelial migration*. In contrast, the majority of Ex-lncRNA/mRNA from the turquoise module and their targeted mRNAs were significantly correlated with *Antigen processing and presentation, Fc γ R-mediated phagocytosis*, and *RIG-I-like receptor signaling pathway* (**Fig. 5**). These results suggest a complex mechanism of a transcriptome regulatory network with distinct roles in modulating immune function between the AW and PW stages of heroin addiction.

**Fig. 5.**
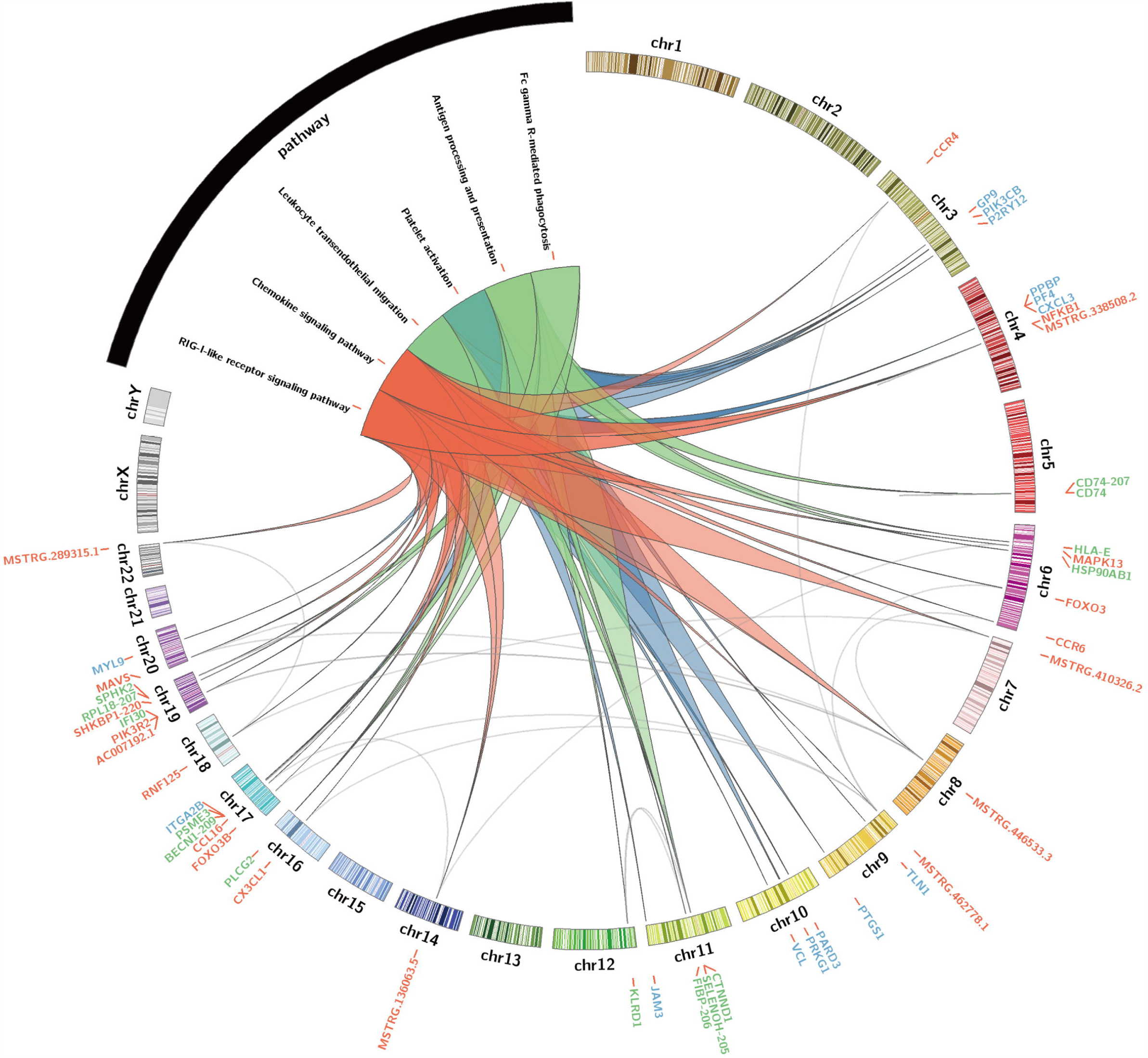
Identification of immune-related lncRNA-mRNA-pathway network. Circos plot showing the top-ranked lncRNA–mRNA-pathway pairs in heroin AW and PW stages. Red, blue and turquoise (green) colors represent the corresponding modules.

## Discussion

Heroin addiction is attracting increasing attention from all elements of the basic, clinical, and public health science-related research communities. Accumulating evidence suggests that the non-coding RNAs involved in the transcriptome regulatory network play significant roles as immune regulators. However, research studies on the mechanism have been rare and there is still much to be learned concerning the molecular basis in the immunosuppression defects frequently observed in heroin addicts. In the present study, using ELISA and high-throughput sequencing technologies, we report the short-term and long-term alterations of circulating cytokines and Ex-lncRNAs/mRNAs in heroin addicts during substance withdrawal. After controlling for various factors (age, BMI, substance dependence history and education), we successfully identified a set of critical lncRNAs and mRNAs in circulating exosomes associated with T_h_1/T_h_2/T_h_17 cytokine balance and switch in heroin addicts.

Over 20 years ago, Govitrapong et al. suggested that heroin addicts in short withdrawal periods experienced decreases in immune system functioning compared to healthy subjects (29). Since then, animal or *ex vivo* models revealed the immunomodulatory effects of heroin and other opioids on cytokine production (5). However, to date, clinical studies investigating immune responses to substance withdrawal and detoxification treatment in human subjects have been limited, and the results have been variable (12, 13). In the present study, it is evident that heroin AW could trigger the T_h_2 cytokines IL-4 and IL-10, and diminish the T_h_1 cytokine IL-2, indicating that heroin has a significant short-term damaging effect on a healthy immune balance, and that the effects of AW are mainly correlated with acute inflammation. Kelschenbach et al. reported that morphine withdrawal contributes to immunosuppression by polarizing T_h_ cells toward the T_h_2 lineage, which is similar to our findings (36). The shift in the T_h_1/T_h_2 cytokine pattern could be reversed in mice by the opioid antagonist naloxone (37). Nevertheless, our results support the concept that the disturbed balance in the T_h_1/T_h_2 ratio contributes to the pathogenesis of heroin withdrawal symptoms. Furthermore, we found a high expression of IL-17A in heroin AW for the first time, indicating that the T_h_17 cells were activated in the early withdrawal period. Zara et al. reported an increase in VEGF in heroin addicts, which is consistent with our data. The level of VEGF increased in heroin AW, indicating the protection of a vessel barrier during withdrawal (38). A prospective study reported that the platelet-lymphocyte ratio was increased in male heroin addicts. In contrast with our data, the level of PDGF-BB decreased in AW. Therefore, it seems that acute inflammatory processes play an important role in the pathophysiology of heroin addicts. However, they are suppressed in AW, perhaps as a self-protection mechanism in acute phase responses (39). Therefore, heroin AW is involved in immune dysregulation, presenting as synchronously activated pro- and anti-inflammation.

In previous studies, the recovery time needed to return to baseline levels varies. Withdrawal-induced immune suppression could last for up to three years in some patients (40). Our study analyzed the alterations of circulating cytokines and the impact of heroin withdrawal on immune system function in a longer time dimension to elucidate the differential cytokine patterns in heroin addicts between the AW and PW groups. Surprisingly, although the dramatically disturbed T_h_1/T_h_2/T_h_17 balance in heroin addicts at the AW stage has been mostly restored to the baseline around one year after the initiation of abstinence, the levels of cytokines of T_h_1 (IL-2, *adj*.*p* = 0.0467; TNF-α, *adj*.*p* = 3.45E-03), T_h_2 (IL-4, *adj*.*p* = 0.06; IL-10, *adj*.*p* = 0.0461), and T_h_17 (IL-17A, *adj*.*p* = 5.41E-03) remained elevated in PW heroin subjects relative to HCs with a statistical significance or a considerable trend toward significance. TNF-*α* is an inflammatory cytokine produced by macrophages/monocytes during acute inflammation and is responsible for various signaling events. An increase in circulating TNF-*α* indicated a chronic inflammation state in heroin addicts even after a year of withdrawal. Meanwhile, the anti-inflammatory cytokine IL-10, which could be produced by virtually all T cells and could serve to antagonize the proinflammatory effects of other cytokines, thereby maintaining the immune balance, remained elevated in PW heroin subjects relative to HCs, suggesting the circulating levels of IL-10, a putative immune biomarker in surveying heroin withdrawal symptoms. Moreover, T_h_17 cells mediate immune responses against extracellular bacteria and fungi, and the dysregulation of IL-17 signaling has been implicated in the pathogenesis of autoimmune diseases and brain disorders. For example, IL-17 serum levels have been linked to morphine withdrawal syndrome in a rat model (41). Our study provides first-in-human evidence indicating that IL-17A may play an important role in the AW and PW stages. In short, we present data showing the dynamics of cytokines in regulating the immune system and inflammatory pathways during different periods of withdrawal. The potential role of these cytokines in immune response suggests that modulating cytokine may be an effective strategy in coping with withdrawal.

In recent years, small, nanosized vesicles (extracellular vesicles) have been identified as a new facet of micro-communication between different organs or cells in the human body, particularly in neurodegenerative and inflammatory diseases (42, 43). These vesicles gain more and more attention and are considered to serve as novel clinical biomarkers and as novel targets for therapeutic drug development (44). By analyzing the differential expression and the regulatory effects of lncRNAs, many studies have identified lncRNAs in association with Alzheimer’s disease, schizophrenia, and depression (45), as well as rheumatoid arthritis and systemic lupus erythematosus (43). However, in the SUD field, the lncRNA biomarker and related mechanistic studies have been lacking. By exploring exosomal RNA contents from peripheral blood, the present study not only identified novel lncRNA biomarkers associated with withdrawal stages in heroin addicts but also revealed withdrawal-stage specific transcriptome regulatory networks.

We also investigated the inflammatory interactome and established associations between dysregulated cytokines and the lncRNA/mRNA co-expression network, which is of great significance to the in-depth study of the influence of heroin addiction and withdrawal on the immune system. To the best of our knowledge, this is the first study to simultaneously characterize the circulating lncRNA and cytokine profiling in plasma samples from heroin addicts and provide evidence that the cytokine-dependent transcriptome network is the functional link between a disturbed immune balance and differential acute and protracted heroin withdrawal symptoms. Moreover, identifying differentially expressed lncRNAs/mRNAs is a commonly used method to explore biomarkers or underlying mechanisms. We identified several novel candidates that may play critical roles in heroin withdrawal symptoms by regulating immune-related pathways. For example, major platelet-derived immune molecules, including the chemokine PF4 and PPBP (**Fig. 5**), were significantly increased at the transcription level in AW heroin addicts. A similar situation was reported in a hypothermic mouse model (46). Considering that the functions of PF4 are complex, in generating pro- and anti-coagulant actions and differentially affecting immune cells (47), we speculate that in response to platelet activation during heroin AW, the platelet-derived PF4 and PPBP transcripts could be encapsulated in circulating exosomes and participate in monocyte recruitment and proinflammatory macrophage differentiation across multiple body sites. Acute inflammation is the protective response of body tissues to harmful stimuli caused by several kinds of tissue injury. The period of acute inflammation was tightly regulated by the immune system; the leukocyte trans-endothelial migration pathway was one of the regulating mechanisms, limiting vascular leakage during leukocytes across the endothelium and promoting leukocyte chemotaxis to the lesion site (48). Consistent with the level of VEGF, the leukocyte trans-endothelial migration pathway was enriched in AW. In addition, trans-endothelial migration of leukocytes involves the spatiotemporal regulation of chemokines (49), hence, *platelet activation, chemokine signaling pathway*, and *leukocyte trans-endothelial migration* point to acute inflammation triggering the process that modulates inflammatory reactions and assists immunocyte migration toward lesions.

Our results show that during heroin PW, the hub lncRNAs/mRNAs CD74, HLA-E, SELENOH-205, RPL18-207, and MSTRG.207040.2 in the turquoise module were significantly upregulated in PW heroin addicts and may also play critical roles in biological processes that are highly correlated with immune responses. CD74 is a cell-surface receptor for the cytokine macrophage migration inhibitory factor and is involved in antigen processing and presentation. MSTRG.207040.2 targets ABCA7 and ARHGAP45 (**Fig. 5**). It has been reported that ABCA7 haplodeficiency disturbs microglial immune responses in the mouse brain, and that ARHGAP45 controls naïve T- and B-cell entry into lymph nodes and T cell progenitor thymus seeding (50). FcγR triggers inflammatory reactions in response to antigen-antibody complexes. It has been reported that the cytokine milieu, such as IL4/10/13, TNF-*α*, regulated FcγR-mediated uptake and inflammation (51). The combination with an *Antigen processing and presentation* enriched pathway suggests that an antigen-related immune response was reactivated by cytokines.

INPP5D, also known as SHIP1, was one of the key factors of the FcγR activated receptor pathway. It is also a multifunction protein. In SHIP1 deficient mice, its loss affects platelet aggregation (52). INPP5D and PLCG2 both participate in the FcγR-mediated pathway. They are also involved in the progression of Alzheimer’s disease, indicating that targeted INPP5D and PLCG2 may have potential in the treatment of heroin addicts (53, 54). Furthermore, MSTRG.136063.7, FIBP-206, MSTRG.7282.1, and IRF7-210 were increased, enriching the RIG-1-like receptor signaling pathway in PW. The activation of RIG-1-like receptor signaling was first identified in heroin addicts. RIG-1 is the important sensor of cytoplasmic viral RNA, and activation of the RIG-1-like receptor was an innate immunity pathway participating in response to viral RNA (55). In sum, it is a signaling pathway for anti-infection in the PW stage. Therefore, although the AW and PW stages are both correlated with immune responses, the molecular inflammatory interactome analyses suggest that unique lncRNA/mRNA-pathway regulatory network results can help to prioritize immune function-related lncRNAs/mRNA and provide a pathway-based view to improve our understanding of their regulatory function in heroin addiction and withdrawal.

A major advantage of this study is the well-balanced clinical characteristics of the two groups of heroin addicts and the HCs, resulting in a true reflection of the relative cytokine quantification in certain withdrawal conditions. Similarly, lncRNA/mRNA in exosomes were enriched and quantitated using a validated protocol (30), offering an opportunity to dissect the correlations between dysregulated cytokine and transcriptome. However, this study has limitations. First, the study was limited by the relatively small sample size. Although we investigated a rather small number of patients, the characteristics of heroin addicts and HCs studied were well balanced, and the two groups of heroin addicts were representative of two typical withdrawal stages (56). This research should be further discussed and validated in large-scale or longitudinal studies. Second, cytokines are also important for brain development and have been implicated in the pathology of a series of brain diseases (42). The impact of heroin or other substances on the brain-immune-axis warrants further investigation. Moreover, the characterization of the lncRNA-mRNA crosstalk network was mainly based on theoretically supported lncRNA-target relationships. Therefore, additional experimental and clinical validation were required. Future studies are necessary to investigate the role of specific lncRNAs and their interactions with relevant mRNAs and to explore the impact of targeted therapeutics on heroin-induced disrupted immunocompetence and associated transcriptome regulatory network changes. In the future, co-administration of anti-inflammatory and anti-infection drugs might be an alternative and more efficient option for heroin addicts during withdrawal. For example, IL-6/7/10/17A inhibitors could be recommended during the entire withdrawal period. However, anti-IL-2 was only used in PW subjects, and the drugs for chemokines/platelet/EGF were only used in AW subjects. Anti-infection agents, especially antiviral infection agents are recommended in PW, depending on the circumstances.

In summary, our study provides a new concept of inflammatory interactome mediated by circulating cytokines and Ex-lncRNA/mRNA, indicating the value of exocellular vesicle encapsulated transcriptome profiling as a biomarker for heroin addiction and withdrawal. These findings open a wide range of future diagnostic and therapeutic options for opioid withdrawal management.

## Data Availability

The clinical data of the patients used to support the findings of this study are included within the article. The original sequencing data of the microRNA used to support the findings of this study are available at GSE172306 of NCBI Sequence Read Archive (SRA).

## Authors contribution statement

ZYZ, HJW, QYP and JHY performed the experiments, researched the data, and wrote the manuscript. ZRX, FRC, YRM, YZZ, YZ, JQY, CC, SYL, YX, HYL, MZ and YQK researched the data and/or helped design experiments. JHY and KHW designed the study, supervised all work, and helped write the manuscript.

## Acknowledgments

This work was supported by grants from the National Natural Science Foundation of China (Grant No. 3171101074, 81860100, 31860306, and 81870458), Science and Technology Department of Yunnan Province (Grant No. 2018DH006, 2018NS0086, 202001AS070004, 202001AV070010) and Yunling Scholar (Grant No. YLXL20170002).

## Figure and table legends

**Supplemental Table 1**. Correlation between Clinical Characteristics and cytokines.

**Supplemental Table 2**. The characteristics of mRNAs with the largest fold change in two groups of heroin addicts versus the group of healthy controls.

**Supplemental Table 3**. The characteristics of lncRNAs with the largest fold change in two groups of heroin addicts versus the group of healthy controls.

**Supplemental Figure 1**. Characterization of exosomes derived from HCs and Heroin addicts peripheral blood. (A) Representative transmission electron micrograph images of exosomes derived from study participants, scale bar□=□200 nm. (B) Representative nanoparticle tracking analysis report of exosomes from study participants. (C) Western blot analysis showing the presence of three common positive exosomal markers (Alix, CD81 and CD63) and one negative exosomal marker (Calnexin) in exosomes isolated from study participants. Ctr positive control, Exo Exosomes experimental group.

