## Supplemental Table 1-3 and Figure 1 for "Integration of molecular inflammatory interactome analyses reveals dynamics of circulating cytokines and extracellular vesicle long non-coding RNAs and mRNAs in heroin addicts during acute and protracted withdrawal"

**Supplemental Figure 1.** Characterization of exosomes derived from HCs and Heroin addicts peripheral blood. (A) Representative transmission electron micrograph images of exosomes derived from study participants, scale bar = 200 nm. (B) Representative nanoparticle tracking analysis report of exosomes from study participants. (C) Western blot analysis showing the presence of three common positive exosomal markers (Alix, CD81 and CD63) and one negative exosomal marker (Calnexin) in exosomes isolated from study participants. Ctr positive control, Exo Exosomes experimental group.

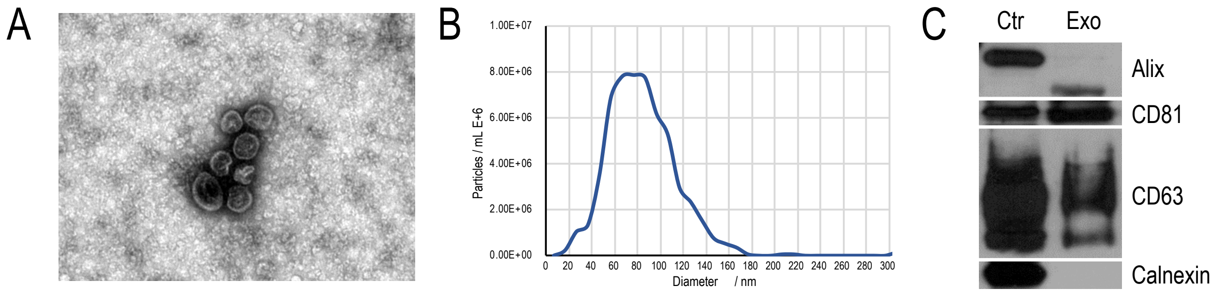

**Supplemental Table 1.** Correlation between Clinical Characteristics and cytokines.

| | **IL-2** | **IL-4** | **IL-5** | **IL-6** | **IL-7** | **IL-8** | **IL-10** | **IL-13** | **IL-17A** | **Basic-FGF** | **G-CSF** | **Eotaxin** | **MCP-1** | **MIP-1α** | **PDGF-BB** | **TNFα** | **VEGF** |
|---|---|---|---|---|---|---|---|---|---|---|---|---|---|---|---|---|---|
| Drug route | 0.23 | -0.02 | 0.16 | -0.10 | 0.13 | **0.34*** | -0.03 | 0.06 | 0.04 | **0.42**** | -0.03 | -0.20 | -0.02 | **0.34*** | **0.36*** | 0.28 | -0.10 |
| Drug type | -0.13 | -0.07 | -0.06 | -0.07 | -0.19 | -0.06 | 0.10 | -0.05 | -0.12 | **-0.34*** | -0.06 | 0.06 | -0.23 | -0.28 | -0.08 | -0.05 | -0.006 |
| Marriage | -0.04 | 0.04 | -0.25 | 0.12 | 0.06 | -0.19 | 0.01 | 0.002 | 0.26 | -0.11 | 0.05 | 0.15 | -0.11 | **-0.36*** | -0.06 | 0.07 | 0.07 |
| Education | 0.11 | 0.27 | 0.31 | 0.005 | **0.36*** | 0.13 | 0.11 | 0.31 | 0.30 | **0.38*** | 0.18 | 0.05 | 0.001 | -0.02 | **0.40*** | 0.11 | 0.06 |
| Age | -0.09 | -0.29 | -0.28 | -0.19 | **-0.34*** | -0.13 | -0.30 | -0.26 | -0.23 | -0.06 | -0.21 | -0.28 | -0.11 | 0.11 | -0.06 | -0.07 | -0.18 |
| BMI | -0.004 | 0.13 | 0.15 | 0.03 | 0.12 | 0.11 | 0.12 | 0.12 | 0.16 | 0.15 | 0.20 | 0.09 | 0.05 | 0.19 | 0.08 | 0.07 | 0.17 |
| Drug years | 0.05 | -0.07 | 0.05 | -0.13 | -0.08 | 0.09 | -0.01 | -0.16 | 0.02 | 0.25 | -0.04 | -0.12 | 0.07 | 0.27 | 0.09 | -0.02 | -0.10 |

| | **TNF-α/IL-4** | **IL-2/IL-4** | **TNF-α/IL-10** | **IL-2/IL-10** | **TNF-α/IL-17A** | **IL-2/IL-17A** | **IL-4/IL-17A** | **IL-10/IL-17A** |
|---|---|---|---|---|---|---|---|---|
| Drug route | 0.13 | 0.15 | 0.11 | 0.07 | 0.10 | 0.21 | -0.05 | 0.04 |
| Drug type | 0.08 | 0.05 | -0.05 | -0.006 | 0.003 | -0.10 | -0.04 | 0.06 |
| Marriage | -0.03 | 0.05 | 0.03 | 0.10 | -0.19 | -0.12 | -0.22 | -0.28 |
| Education | -0.15 | -0.009 | 0.007 | 0.01 | 0.01 | 0.02 | 003 | -0.05 |
| Age | 0.12 | -0.008 | 0.05 | 0.08 | 0.19 | -0.02 | -0.14 | -0.24 |
| BMI | -0.10 | -0.11 | -0.14 | -0.16 | -0.03 | 0.06 | 0.03 | -0.02 |
| Drug years | -0.21 | -0.007 | **-0.34*** | **-0.33*** | -0.11 | 0.03 | -0.07 | -0.002 |

* $p < 0.05$, the first four rows are Spearman's correlation, and the last three rows are Pearson's correlation.

**Supplemental Table 2.** The characteristics of mRNAs with the largest fold change in two groups of heroin addicts versus the group of healthy controls.

| Gene Symbol | AW vs HC | p | Gene Symbol | PW vs AW | p | Gene Symbol | PW vs HC | p |
|---|---|---|---|---|---|---|---|---|
| **CD3D** | **-2.814721488** | **0.0018519** | **MAFA** | **-3.743466027** | **0.0002252** | AL133352.1 | -2.12071603 | 0.01348041 |
| **CLC** | **-2.108972099** | **0.03589759** | **H2AW** | **-2.775148463** | **0.00044147** | **H2AW** | **-2.033832622** | **0.00305922** |
| KLRD1 | -1.863037981 | 0.00061356 | EGF | -2.174140003 | 0.00083505 | **CLC** | **-1.88378074** | **0.02476634** |
| GPR18 | -1.852498563 | 0.00053102 | AZU1 | -2.170193417 | 0.00544115 | PARL | -1.883129529 | 0.00877705 |
| KLRC4-KLRK1 | -1.767054365 | 0.01112618 | MFAP3L | -2.090923847 | 0.00102643 | **CCNB2** | **-1.853662165** | **0.00149792** |
| CD3G | -1.746148559 | 0.00020203 | DEFA4 | -2.049861319 | 0.01483743 | PFDN6 | -1.801627654 | 0.00946389 |
| FHDC1 | -1.734391829 | 0.0477427 | WBP1 | -2.037150089 | 0.00392813 | CCDC77 | -1.79289857 | 0.00156027 |
| **SH2D1A** | **-1.601420582** | **0.00188209** | MT1X | -2.012413235 | 0.009491 | PTTG1 | -1.785036663 | 0.01810236 |
| ARL4C | -1.598837066 | 0.03290972 | **CCNB2** | **-1.980150792** | **0.00322177** | RHD | -1.774578734 | 0.00355547 |
| C12orf42 | -1.588090399 | 0.00042015 | **ACRBP** | **-1.961879675** | **0.01533507** | KLRC3 | -1.75371719 | 0.00921878 |
| HOMER2 | 1.912408335 | 0.0000301 | AAMDC | 1.461926341 | 0.02179978 | NFIC | 1.39700295 | 0.00586449 |
| SPX | 1.96782632 | 0.0000153 | DNASE1L3 | 1.462829525 | 0.00779191 | JAM3 | 1.397697051 | 0.04214788 |
| STX3 | 1.989037506 | 0.00034286 | NDUFA7 | 1.47580543 | 0.01005825 | C11orf24 | 1.430208502 | 0.00214423 |
| S100P | 1.990360689 | 0.02687818 | **SH2D1A** | **1.538772383** | **0.00028682** | IGFBP4 | 1.540958939 | 0.00323664 |
| GP9 | 2.089445713 | 0.02568544 | HAUS2 | 1.583187388 | 0.00851377 | TEAD4 | 1.563891757 | 0.00802384 |
| **ACRBP** | **2.131116143** | **0.01546211** | SULT1A3 | 1.666447798 | 0.00173937 | ADIRF | 1.567954384 | 0.01820304 |
| SNCG | 2.16056009 | 0.04164684 | TRIM34 | 1.675228496 | 0.03805604 | SMIM24 | 1.596202464 | 0.00427664 |
| **NRIP1** | **2.180488799** | **0.00066636** | INO80B-WBP1 | 1.87888765 | 0.00413145 | NDRG2 | 1.596480132 | 0.0009078 |
| **MAFA** | **2.205464527** | **0.0148821** | CCL21 | 2.109545567 | 0.00082991 | **NRIP1** | **1.86516547** | **0.00095734** |
| **C19orf33** | **3.020128756** | **0.00473678** | **CD3D** | **2.24438949** | **0.00531615** | **C19orf33** | **2.054818438** | **0.02411904** |

**Supplemental Table 3.** The characteristics of lncRNAs with the largest fold change in two groups of heroin addicts versus the group of healthy controls.

| Gene Symbol | Targeted gene | | AW vs HC | p | Gene Symbol | Targeted gene | | PW vs AW | p | Gene Symbol | Targeted gene | | PW vs HC | p |
|---|---|---|---|---|---|---|---|---|---|---|---|---|---|---|
| MSTRG.237243.6 | CNGA3;INPP4A | cis | -3.159 | 2.81E-05 | **MSTRG.136063.5** | RPS29;LRR1;RPL36AL;MGAT2;DNAAF2;POLE2;KLHDC1;KLHDC2;NEMF;ARF6 | cis | -5.7 | 0.004551116 | MSTRG.192806.4 | SEPTIN9 | cis | -3.7229 | 4.62E-05 |
| MSTRG.116065.8 | FBRSL1;LRCOL1;P2RX2;POLE;PXMP2 | cis | -2.857 | 0.001437763 | MSTRG.410326.2 | SDK1 | cis | -3.45 | 6.43E-06 | **EIF4A1-207** | EIF4A1 | cis | -3.5609 | 0.00436721 |
| MSTRG.274372.465 | | | -2.853 | 0.000464466 | MSTRG.274376.48 | | | -3.05 | 0.047622607 | MSTRG.422578.3 | SRRM3;HSPB1;YWHAG | cis | -3.2598 | 0.00291457 |
| **EIF4A1-207** | EIF4A1 | cis | -2.63 | 0.024316359 | MSTRG.22804.2 | TENT5C | cis | -2.87 | 0.000210477 | **MSTRG.35400.2** | SLC26A9;RAB7B;CTSE;RHEX | cis | -2.8207 | 0.00036048 |
| MSTRG.274378.7 | | | -2.518 | 0.045336802 | MSTRG.409997.2 | INTS1;MAFK;TMEM184A;PSMG3 | cis | -2.72 | 0.000541643 | MSTRG.45110.2 | SFMBT2 | cis | -2.7455 | 0.00684349 |
| **RPL23-210** | RPL23 | cis | -2.482 | 0.02131207 | MSTRG.279447.7 | PCP4;DSCAM | cis | -2.67 | 0.015447039 | EEF1G-203 | EEF1G | cis | -2.7123 | 0.02631233 |
| MSTRG.182620.2 | MAP2K3;KCNJ12 | cis | -2.477 | 0.017450841 | MSTRG.101316.2 | KRT2;KRT1;KRT77;KRT76;KRT3;KRT4;KRT79;KRT78 | cis | -2.63 | 0.000223037 | MSTRG.78659.2 | SCYL1;LTBP3;ZNRD2;FAM89B;EHBP1L1;KCNK7;MAP3K11;PCNX3 | cis | -2.6369 | 0.00584422 |
| OXA1L-207 | OXA1L | cis | -2.324 | 0.019862464 | MSTRG.25448.2 | TCHH;RPTN;HRNR | cis | -2.6 | 0.001560047 | MSTRG.324272.2 | SH3TC1;HTRA3 | cis | -2.588 | 0.00644348 |

| | | | | | | | | | | | | | | |
|---|---|---|---|---|---|---|---|---|---|---|---|---|---|---|
| DNAJC7-204 | DNAJ7C | cis | -2.262 | 0.014598663 | **MSTRG.148661.3** | GOLGA8K | cis | -2.57 | 0.002124975 | MSTRG.133950.3 | EGLN3;SPTSSA;EAPP;SNX6 | cis | -2.5127 | 0.03615019 |
| MSTRG.366349.7 | LNPEP;LIX1;RIOK2 | cis | -2.241 | 0.014672079 | **MSTRG.35400.2** | SLC26A9;CTSE;RHEX | cis | -2.52 | 0.002283715 | MSTRG.426186.3 | IFT22;COL26A1 | cis | -2.4914 | 0.00183151 |
| MSTRG.468402.2 | | | 2.1861 | 0.00239438 | MSTRG.212951.2 | TSHZ3 | cis | 2.078 | 0.021340914 | **MYL6-216** | MYL6 | cis | 1.97872 | 0.00416429 |
| ACRBP-206 | ACRBP | cis | 2.1868 | 0.010576816 | ARHGAP15-209 | ARHGAP15 | cis | 2.082 | 0.015162068 | CXXC1-217 | CXXC1 | cis | 2.06058 | 0.00459133 |
| NRIP1-205 | NRIP1 | cis | 2.1925 | 0.006409114 | **MYL6-216** | MYL6 | cis | 2.144 | 0.001835834 | CNDP2-222 | CNDP2 | cis | 2.08329 | 0.02874254 |
| **MSTRG.148661.3** | GOLGA8K | cis | 2.2633 | 0.005782681 | **MSTRG.136063.4** | RPS29;LRR1;RPL36AL;MGAT2;DNAAF2;POLE2;KLHDC1;KLHDC2;NEMF;ARF6 | cis | 2.204 | 0.04070416 | MSTRG.460211.2 | | | 2.1494 | 0.00790498 |
| STX3-205 | STX3 | cis | 2.3383 | 0.001590537 | NAP1L1-210 | NAP1L1 | cis | 2.23 | 0.001886827 | MKRN1-209 | MKRN1 | cis | 2.26244 | 0.02992848 |
| GATAD2A-209 | GATAD2A | cis | 2.341 | 0.002138832 | **RPL18-207** | RPL18 | cis | 2.316 | 6.29E-05 | **RPL8-214** | RPL8 | cis | 2.3918 | 0.00039393 |
| MSTRG.81169.2 | LRRC32 | cis | 2.6048 | 0.000382834 | RPLP2-208 | RPLP2 | cis | 2.509 | 0.046068913 | **RPL18-207** | RPL18 | cis | 2.72427 | 5.92E-07 |
| PSME1-209 | PSME1 | cis | 3.334 | 0.012774505 | **RPL23-210** | RPL23 | cis | 2.51 | 0.004516473 | MSTRG.274376.37 | | | 3.36027 | 0.01270372 |
| MSTRG.136063.8 | RPS29;LRR1;RPL36AL;MGAT2;DNAAF2;POLE2;KLHDC1;KLHDC2;NEMF;ARF6 | cis | 3.6516 | 0.034533166 | **RPL8-214** | RPL8 | cis | 2.795 | 3.70E-07 | MSTRG.136063.7 | RPS29;LRR1;RPL36AL;MGAT2;DNAAF2;POLE2;KLHDC1;KLHDC2;NEMF;ARF6;VCPKMT;SOS2;L2HGDH | cis | 3.72424 | 0.00396484 |

| | | | | | | | | | | | | | |
|---|---|---|---|---|---|---|---|---|---|---|---|---|---|
| **MSTRG.136063.5** | RPS29;LRR1;RPL36AL;MGAT2;DNAAF2;POLE2;KLHDC1;KLHDC2;NEMF;ARF6 | cis | 3.9088 | 0.034411535 | RIOK3-205 | RIOK3 | cis | 2.801 | 0.043527386 | **MSTRG.136063.4** | RPS29;LRR1;RPL36AL;MGAT2;DNAAF2;POLE2;KLHDC1;KLHDC2;NEMF;ARF6;VCPKMT;SOS2;L2HGDH | cis | 4.06835 | 0.01851068 |
